# Human Resource Challenges in Leprosy Control: A Cross-Sectional Study in Southwest border area of China

**DOI:** 10.1101/2025.06.09.25329245

**Authors:** Ruifang Song, Yong Shen, Fuying Guo, Qing Zhen, Shuo Kou, Shun Zha, Xiaojun Yu, Zhuo Li, Shanshan Song, Jiaxin Hao, Yiting Zhang, Yingtong Wang, Tian Ma, Tiejun Shui, Xiangyu Yan, Weijia Zhao

## Abstract

**Background:** Leprosy remains a neglected tropical disease and major global public health challenge, particularly in developing countries with severe healthcare workforce shortages hindering control. The southwestern border, represented by Yunnan Province, is a core endemic area. A comprehensive assessment of its leprosy workforce is critical for achieving elimination.

**Methods/Results:** In October 2024, a cross-sectional census survey evaluated leprosy prevention personnel across all 126 counties in Yunnan Province, China. These counties were categorized into high (I), moderate (II), and low (III) endemicity areas based on their leprosy burden. Distinct workforce patterns emerged among 423 personnel, Category I areas exhibited the highest proportion of personnel aged 30–39 years (37.5%), along with the most educated workforce (78.5% holding a bachelor’s degree or higher) and the highest full-time employment rate (52.5%). Category II areas featured the oldest personnel profile (35.0% aged ≥ 50 years), highest continuing education participation (78.8%), and strongest prescription protocol awareness (72.5%). In contrast, Category III areas had the highest proportion under 30 years (20.5%), yet the lowest education levels (69.9% above bachelor’s degreer), lowest full-time rate (38.4%), highest compensation dissatisfaction (38.4% “below average”), and lowest intention to leave (8.2%). Pervasive workforce aging existed (at least 30% of personnel ≥ 50 years in Category II and III) and widespread technical gaps (>75% Category III areas lacked essential lab skills). Part-time staffing was common (47.5%-61.6% across categories). Despite >95% of personnel rating compensation as “average”, compensation and career challenges were most acute in Categories I and II.

**Conclusions:** The Yunnan leprosy workforce shows strengths, notably high continuing education participation (78.5%) and balanced clinical/preventive staffing. However, it faces significant challenges, workforce aging, shortage of highly qualified personnel, and limited lab capacity. Urgent intervention measures are needed to revitalize the workforce, enhance training, and strategically allocate resources to expedite the achievement of leprosy elimination.

## Introduction

Leprosy, a chronic infectious disease caused by *Mycobacterium leprae*, has seen a 97% reduction in annual reported cases worldwide since the World Health Organization (WHO) introduced multidrug therapy (MDT) in 1985[1]. However, 2023 data reveal that low- and middle-income countries (LMICs) continue to bear 90% of the global disease burden.This paradox stems from structural imbalances in healthcare resource allocation[2]. In 2023, the WHO highlighted critical gaps in leprosy-endemic regions, Africa and Southeast Asia have a leprosy specialist density of only 0.2 per 100,000 population, and 60% of grassroots healthcare facilities lack standardized diagnostic tools, contributing to a case detection delay rate of 28.3%[2]. For instance, in rural India, healthcare worker density is less than one-third of that in urban areas, increasing the risk of irreversible nerve damage by 2.3-fold[3]. Similarly, Brazilian studies reveal that only 34% of primary care physicians master early diagnostic skills for leprosy. These findings underscore the global bottleneck of workforce shortages and weak surveillance capacity in achieving leprosy elimination[4].

China has significantly reduced leprosy prevalence through national interventions[5–7]. However, Yunnan Province, the country’s hyperendemic epicenter, demands urgent attention due to its unique challenges[8]. On the one hand, for the epidemiological burden,Yunnan contributes over 25% of national new cases, with elimination progress lagging behind the national average by more than a decade[6, 8, 9]. On the other hand, Yunnan Province faces geographical and sociocultural complexity, located in southwestern China (97.31 ° E – 106.11 ° E, 21.80 ° N – 29.15 ° N), spanning 394,000 km ² , of which 94% is mountainous terrain, posing challenges for leprosy control efforts[10, 11]. Bordering Myanmar, Laos, and Vietnam, it serves as a key hub for public health cooperation in the Greater Mekong Subregion (GMS)[12, 13](Fig 1). Rugged landscapes and linguistic diversity among 26 ethnic groups hinder healthcare accessibility. Surveillance data reveal a case detection delay rate of 32.1%, and 12.3% of patients develop irreversible disabilities, which is 65% higher than the national average. This situation traps patients in a “treatment-disability-poverty” cycle.

**Fig 1.**
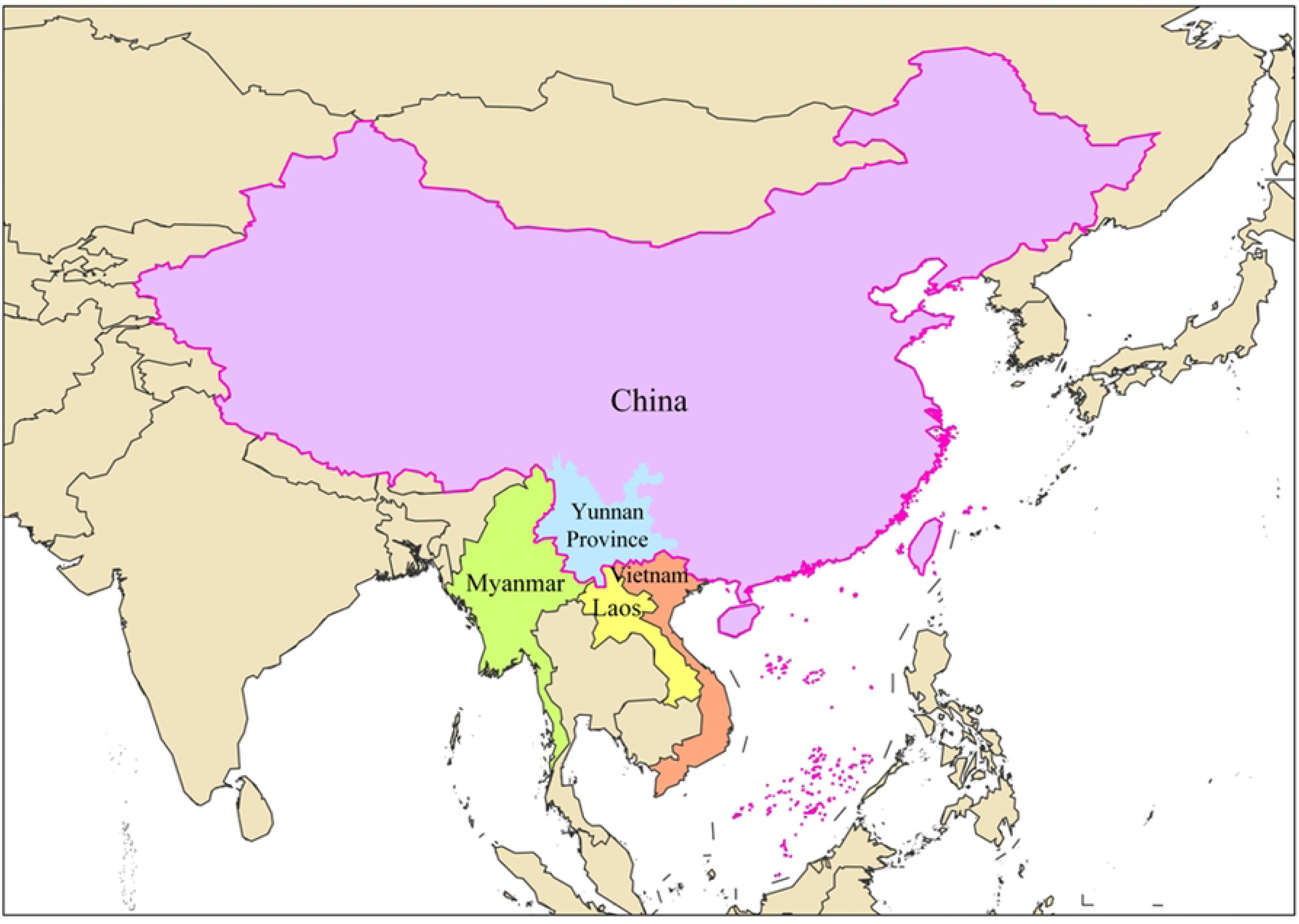
The unique geographical location of Yunnan within China’s southwestern border region

International experience has shown that leprosy elimination is highly dependent on the size and specialized capacity of human resources for health at the grass-roots level. Currently, high-burden countries around the world are generally facing a triple dilemma[14]. First, there is a lack of pathogenic data-sharing mechanisms. Only 41% of health posts in rural areas of Africa are equipped with skin smear testing equipment. Second, there is a disconnection in the training system for medical personnel[15]. According to the WHO 2023 report, only 45% of leprosy prevention and treatment personnel globally have received systematic skills training[16]. Third, the coverage of the surveillance network is insufficient. The misdiagnosis rate of leprosy in health institutions in mountainous areas of Indonesia is as high as 37%[17, 18]. This dilemma is particularly prominent in cross-border disease transmission hotspots where countries in the Greater Mekong Subregion (GMS) have difficulty in realizing coordinated cross-border tracking of cases due to uneven distribution of medical resources[17, 19]. Yunnan Province, as a core node in the region, will have more than 12 million cross-border population movements in 2021, but the intensity of CDC staffing in border counties is only 57% of that in inland areas[20].

To address these issues, Yunnan initiated the *Three-Year Action Plan for High-Quality Health Development* in 2024, aiming to achieve county-level elimination[21]. However, baseline evaluations have revealed several structural contradictions[22]. There is a mismatch between the policy mandates for county-wide coverage and the existing workforce shortages. Disparities also exist between the needs for precision control and the homogenized nature of grassroots services. Furthermore, the multi-sector mechanisms for disability interventions, which involve civil affairs and education systems, remain uncoordinated. Despite these efforts, the health workforce and capacity in the professional field of leprosy prevention and control have not yet been comprehensively evaluated.

The aim of this study is to provide the first comprehensive assessment of Yunnan’s leprosy control workforce, evaluating workforce structure, human resource distribution, laboratory capacity, professional composition, and disciplinary balance. Through analysis of epidemiological gradient characteristics, the study identifies targeted strategies for optimizing human capital in high-burden regions and provides insights applicable to global elimination efforts.

## Materials and Methods

### Study Design

We conducted a census-based cross-sectional study systematically investigating leprosy prevention and control personnel across 129 counties (districts) within 16 prefecture-level divisions in Yunnan Province, China, from October 1 to October 31, 2024, with structured questionnaire surveys administered on-site during field investigations. The survey encompassed all levels of the provincial prevention and control system, including staff at the prefectural/municipal level and county/district level. County-level units were classified into three categories based on the *China’s National Leprosy Elimination Plan (2011-2020)* and the *Yunnan Provincial Implementation Plan for Leprosy Elimination*, combined with local epidemiological characteristics: Category I (High-endemic areas), Category II (Moderate-endemic areas), and Category III (Low-endemic areas) (Fig 2) (Supplementary Table 1).

**Fig 2.**
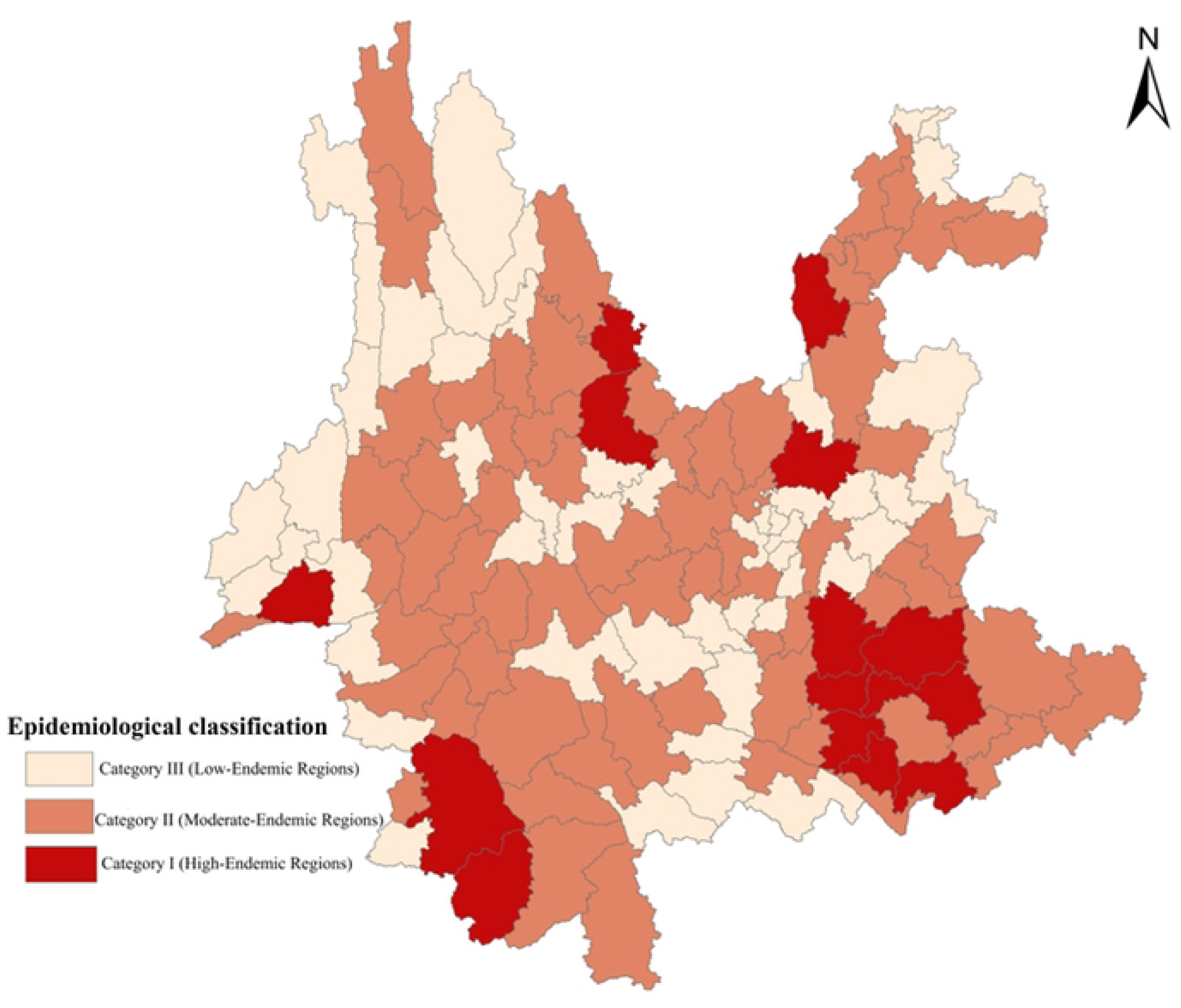
Spatial Distribution of Epidemiological Risk Levels in Yunnan Province

### Study Participants

This study implemented dual coverage of institutional and personnel censuses across all leprosy prevention and control institutions in Yunnan Province. At the institutional level, the survey encompassed all Centers for Disease Control and Prevention (CDC) institutions, designated hospitals, and grassroots healthcare facilities engaged in leprosy control. At the personnel level, the study included all on-duty professionals involved in leprosy prevention and control, categorized into four roles: administrative staff, clinical physicians, laboratory technicians, and nursing/rehabilitation personnel. Temporary or non-active personnel (e.g., those on secondment or training) were excluded. This study uses a dual classification framework, on the one hand, personnel are divided into provincial or county-level staff. On the other hand, county-level administrative units are classified into category I (high-prevalence) areas, category II (medium-prevalence) areas , and category III (low-prevalence) areas based on the prevalence of leprosy.

### Ethical Approval

The study was approved by the Medical Ethics Committee of the Yunnan Provincial Center for Disease Control and Prevention (Approval No. 2024-70). All participants provided written informed consent, and data were anonymized with sensitive information aggregated to ensure confidentiality during analysis.

### Statistical Analysis

Data were independently double-entered into EpiData 3.1 with consistency validation procedures. Statistical analyses were performed in SPSS 24.0, including descriptive analyses of demographic and occupational variables. Spatial distribution maps were generated using ArcMap 10.8. Categorical variables were reported as absolute frequencies or proportions. chi-square test or Fisher exact test, were employed to assess associations between variables, with statistical significance set at α = 0.05.

## Results

### Between Prefecture and County-Level Leprosy Control Workforce

At present, most of the leprosy prevention and control personnel in Yunnan Province are concentrated in Wenshan, followed by Pu’er and Dali (Fig 3).

**Fig 3.**
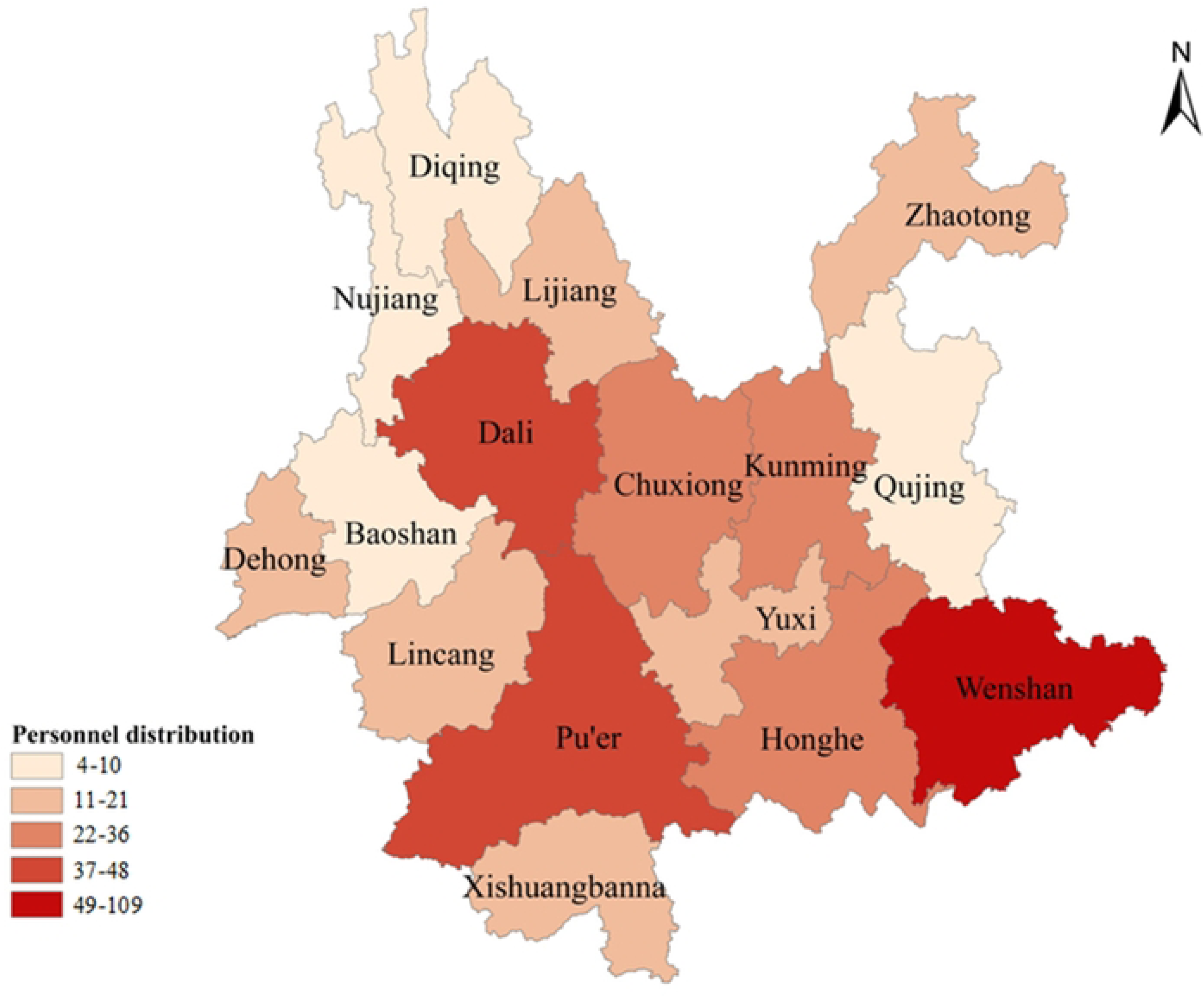
Distribution of leprosy prevention and control personnel inYunnan Province

The study identified systematic variations in workforce characteristics between administrative levels. At the prefecture level, personnel aged 30–39 years comprised 40.3% of the workforce, a proportion significantly higher than the 26.4% observed at the county level. Conversely, county-level staff demonstrated greater representation of individuals aged 50–59 years (31.5%) compared to the prefecture-level workforce (19.4%) (χ² = 12.839, p = 0.012). Ethnic minority composition showed a distinct pattern, constituting 37.6% of county-level personnel versus 23.9% at the prefecture level (χ² = 4.665, p = 0.031). Educational attainment diverged markedly, with 76.1% of prefecture-level professionals holding bachelor’s degrees compared to 60.4% at the county level (χ² = 44.479, p < 0.001) (Table 1).

Professional qualifications exhibited hierarchical stratification. Prefecture-level personnel predominated in public health expertise (47.8%) and public health practicing physician certifications (71.8%), whereas county-level counterparts focused on clinical medicine specializations (48.3%) and clinical practicing physician certifications (61.5%) (specialization χ² = 25.336, p < 0.001, certification χ² = 26.330, p < 0.001). Full-time employment rates were higher at the prefecture level (62.7%) relative to county-level institutions (48.0%) (χ² = 4.843, p = 0.028), though resignation intentions were more prevalent among county staff (10.7%) than prefecture-level personnel (4.5%).

Capacity-building metrics revealed 88.1% training participation at prefecture level and 86.8% at county level, with continuing education completion rates of 85.1% and 74.4% respectively. Despite these efforts, critical diagnostic infrastructure gaps persisted—only four prefectures maintained pathological diagnostic capacity, with 75% of primary institutions lacking this capability (Fig 4). Regarding self-reported compensation levels, 91% of prefectural-level respondents and 95.3% of county-level respondents rated their compensation as “below average”. (Table 1).

**Fig 4.**
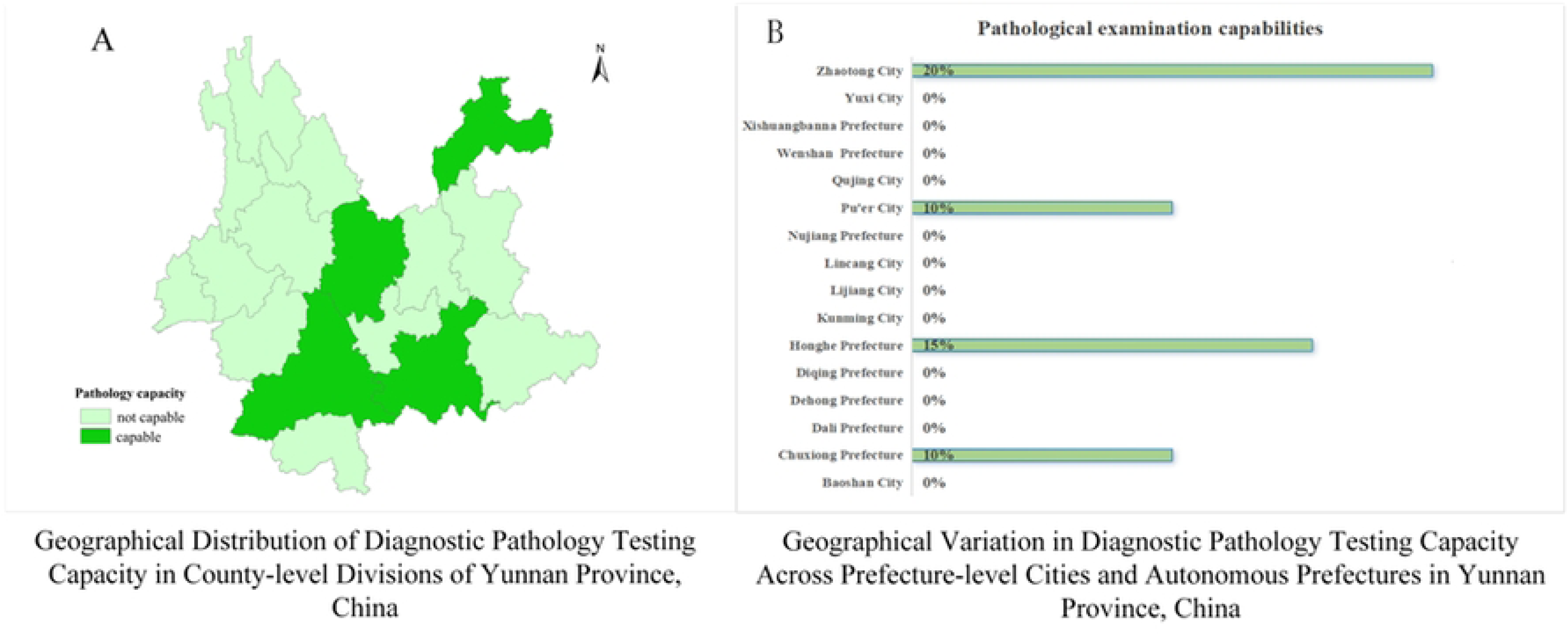
Pathological Examination Capabilities Across Prefectures and Cities in Yunnan Province

### Comparison of Different Endemic Areas in County-Level

The study revealed distinct workforce patterns across leprosy risk stratification areas. Category I areas demonstrated a higher proportion of younger professionals aged 30–39 years (37.5%) compared to Category II (23.6%) and Category III (21.9%), while aged 50–59 years were more prevalent in Category II (35.0%) and III (32.9%) than in Category I (21.3%) (Table 2). Ethnic minority representation varied geographically, with Category I (41.2%) and III (38.4%) areas showing greater diversity compared to Category II (36.0%).

Professional qualifications exhibited systematic stratification. Category III areas reported the highest bachelor’s degree attainment (69.9%), surpassing both Category I (58.8%) and II (57.6%). Clinical medicine specialization dominated in Category I (46.3%) and II (55.2%) areas, correlating with higher clinical practicing physician certification rates (72.7% and 65.4%, respectively). In contrast, Category III areas showed stronger public health orientation (45.2% specialization, 51.3% public health certifications). Full-time employment rates were lowest in Category III (38.4%) relative to Category I (52.5%) and II (49.8%) (Table 2).

Operational capacities displayed paradoxical trends. Training participation exceeded 83% across all areas, yet continuing education completion in Category III (54.8%) lagged behind Category I (78.5%) and II (78.8%). Prescription authority awareness was higher in Category I (72.5%) than in other regions (50.2–52.1%). While all 16 prefectures in Yunnan Province have established basic diagnostic capacity for skin tissue fluid smear examinations, 37% of county-level divisions (48/129) remain unequipped, despite exemplary coverage in four prefectures achieving full implementation (Fig 5). In the three types of areas, more than 95% of personnel reported that their salary levels were average or below average, but their intention to resign was about 10% (Table 2).

**Fig 5.**
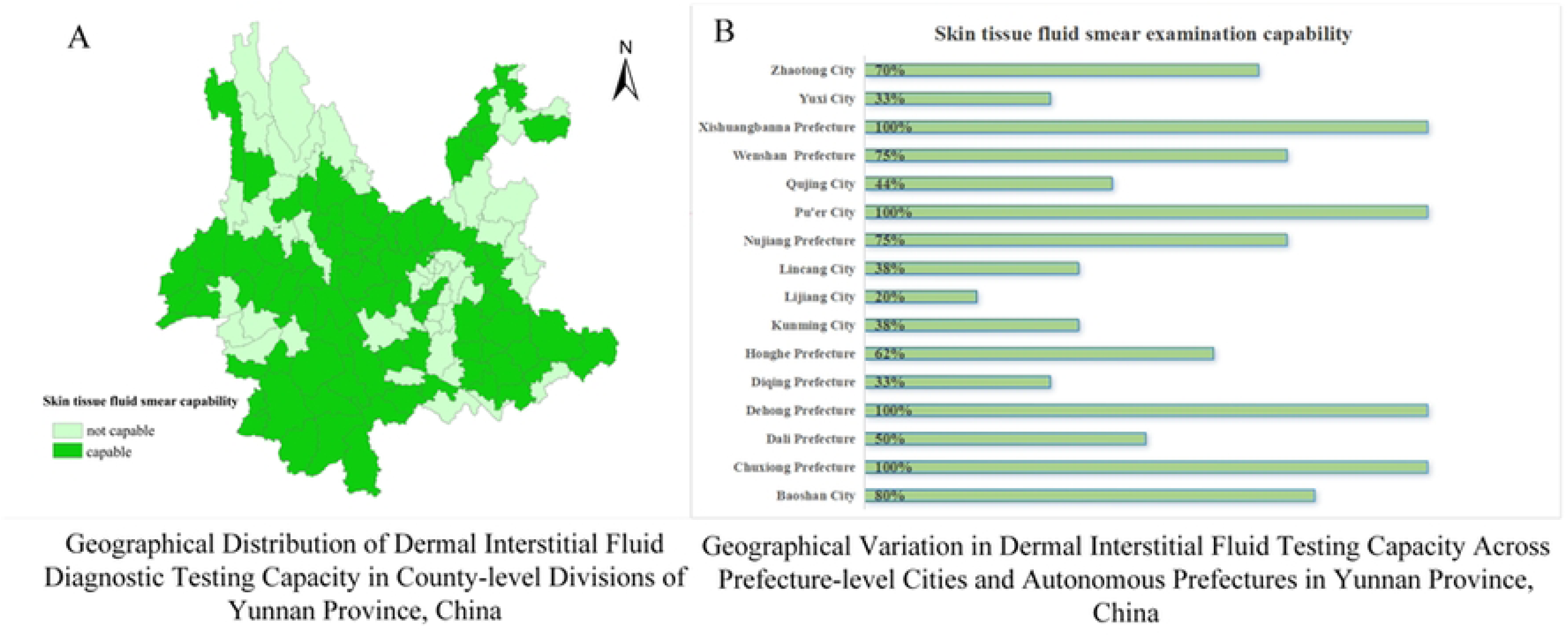
Spatial Distribution of Skin Tissue Fluid Smear Diagnostic Capacity in Yunnan Province

## Discussion

The cross-sectional study on the current status of leprosy prevention workforce development in Yunnan Province reveals multidimensional challenges in grassroots prevention systems. The findings align with global research on human resources for leprosy control. This analysis, focusing on age structure, institutional management, professional composition, staffing stability, turnover intention, and technical capacity, provides critical evidence for optimizing resource allocation in leprosy prevention.

The study highlights significant workforce aging in Yunnan, particularly in Category II and III regions, where personnel aged 50–59 account for 35.0% and 32.9%, respectively, while those under 30 represent less than 20%. Similar aging trends have been observed in other regions. For example, In sub-Saharan Africa, over 30% of healthcare workers are aged 50 years or older[23]. In India, rural areas face significant challenges in attracting and retaining younger health professionals, leading to an aging and overburdened workforce[24]. Aging workforces risk knowledge transfer gaps and reduced adaptability to emerging technologies (e.g., digital case management systems)[25].

Leprosy control personnel exhibited a fragmented institutional distribution in Yunnan Province, spanning CDC facilities (47.5%–98.6%), dermatology stations, and general hospitals, while part-time staffing reached 47.5%–61.6% across categories. This institutional fragmentation, compounded by legal restrictions on diagnostic authority under China’s Practicing Physician Law, has resulted in diagnostic authority gaps among CDC personnel. Similar systemic misalignment has been documented in India’s leprosy control system[26], underscoring how disjointed interdepartmental collaboration undermines operational efficiency. Establishing cross-sector coordination mechanisms and clarifying institutional responsibilities are critical to optimizing prevention outcomes.

In Category III regions, over 45% of personnel perceive their compensation as “average,” and continuing education participation drops to 54.8%—significantly lower than in high-endemic regions (78.5%). Lack of competitiveness in remuneration and narrow scope for career development, which may lead to the phenomenon of “passive job retention”.These issues coincide with a study of burnout among health workers in 107 institutions in 18 provinces in the leprosy region of China[28]. Enhancing salary structures, career advancement opportunities, and training investments are crucial to improving job satisfaction and retention.

Slit-skin smear coverage (62.8%) and pathological diagnostic capacity (4.7%) in Yunnan lag behind Southeast Asian averages (85% and 12%, respectively). With global Mycobacterium leprae drug resistance rising from 1.2% (2010) to 3.8% (2022)[29], strengthening laboratory networks and antimicrobial resistance surveillance is urgent[30].

Additionally, Yunnan’s leprosy control system demonstrates several unique strengths that are adaptable on a global scale. First, Yunnan leads in continuing education participation, with rates in high-endemic regions reaching 78.5%, which exceed those in Brazil (58%) and India (52%)[31]. Second, Yunnan has strong diagnostic coverage, with slit-skin smear availability at 62.8%, surpassing rates in Cambodia (35.4%) and Laos (28.1%)[32]. Third, Yunnan demonstrates adaptive professional allocation, with a clinical focus in high-endemic areas (46.3%) and a public health emphasis in low-endemic zones (45.2%). This aligns with the WHO’s precision control principles and has reduced misdiagnosis rates by 34% compared to Ethiopia’s clinical-dominated model[33].

## Conclusion

In conclusion, this pioneering study constitutes the first comprehensive elucidation of multilevel structural barriers to human resource optimization in Yunnan’s leprosy control system, establishing an empirical foundation for evidence-based elimination strategies aligned with the WHO’s 2030 leprosy eradication milestones and China’s Healthy Yunnan 2030 roadmap for zero transmission. We conducted a thorough assessment of leprosy control personnel across multiple dimensions, including demographics, organizational structure, professional background and qualifications, employment patterns, technical competencies, professional development, perceived income satisfaction, and turnover intention. The results indicate that Yunnan has strengths in continuing education participation, leprosy-specific training, and the distribution of clinical and preventive professionals across different endemic areas. However, several issues persist, including workforce aging, a shortage of highly educated talent, insufficient integration of medical and preventive services, a high proportion of part-time staff, and limited technical capacity. It is recommended that public health authorities implement integrated strategies combining youth recruitment programs, cross-sector collaboration models, and targeted policy support to enhance workforce retention and technical proficiency. Future research should focus on prospective cohort studies to evaluate the long-term effectiveness of these interventions and further strengthen global leprosy elimination strategies.

## Limitations

First, self-reported measures of compensation satisfaction, resignation intentions, and training participation may be influenced by social desirability bias and recall inaccuracies. Second, diagnostic capacity assessment relied exclusively on equipment availability (e.g., skin smear tools) without evaluating operational proficiency or diagnostic accuracy, potentially overestimating technical capabilities.

## Data Availability

The datasets generated and analyzed during this study are included in the published article. Additional underlying data are available from the corresponding author upon reasonable request.

## Acknowledgment

We gratefully thank the staff members of the Center for Disease Control and Prevention of Yunnan for their assistance in the data collection, and validation.

